# A curated census of pathogenic and likely pathogenic UTR variants and evaluation of deep learning models for variant effect prediction

**DOI:** 10.1101/2023.07.10.23292474

**Authors:** Emma Bohn, Tammy Lau, Omar Wagih, Tehmina Masud, Daniele Merico

**Author notes:** These authors contributed equally to this work and share first authorship (Emma Bohn, Tammy Lau). Correspondence: Dr. Tehmina Masud, Deep Genomics, Toronto, ON, Canada, Dr. Daniele Merico, Vevo Therapeutics, San Francisco, CA, USA (current affiliation).

## Abstract

Variants in 5’ and 3’ untranslated regions (UTR) contribute to rare disease. While predictive algorithms to assist in classifying pathogenicity can potentially be highly valuable, the utility of these tools is often unclear, as it depends on carefully selected training and validation conditions. To address this, we developed a high-confidence set of pathogenic (P) and likely pathogenic (LP) variants and assessed deep learning (DL) models for predicting their molecular effect. 3’ and 5’ UTR variants documented as P or LP (P/LP) were obtained from ClinVar and refined by reviewing the annotated variant effect and reassessing evidence of pathogenicity following published guidelines. Prediction scores from sequence-based DL models were compared between three groups: P/LP variants acting though the mechanism for which the model was designed (model-matched), those operating through other mechanisms (model-mismatched), and putative benign variants. PhyloP was used to compare conservation scores between P/LP and putative benign variants. 295 3’ and 188 5’ UTR variants were obtained from ClinVar, of which 26 3’ and 68 5’ UTR variants were classified as P/LP. Predictions by DL models achieved statistically-significant differences when comparing model-matched P/LP variants to both putative benign variants and model-mismatched P/LP variants, as well as when comparing all P/LP variants to putative benign variants. PhyloP conservation scores were significantly higher among P/LP compared to putative benign variants for both the 3’ and 5’ UTR. In conclusion, we present a high-confidence set of P/LP 3’ and 5’ UTR variants spanning a range of mechanisms and supported by detailed pathogenicity and molecular mechanism evidence curation. Predictions from DL models further substantiate these classifications. These datasets will support further development and validation of DL algorithms designed to predict the functional impact of variants that may be implicated in rare disease.

## 1 Introduction

As the diagnostic utility of whole genome sequencing (WGS) in rare disease populations is increasingly documented (<u>Clark et al., 2018</u>; <u>Stavropoulos et al., 2016</u>), there is a growing appreciation for the direct implication of non-coding variation in heritable disease (<u>French & Edwards, 2020</u>). Clinically relevant variants have been identified in a range of functional elements residing in non-coding regions, including 5’ and 3’ untranslated regions (UTRs) (<u>Steri et al., 2018</u>; <u>Chatterjee & Pal, 2012</u>). These regions flank the coding sequence, and play an important role in mRNA stability, translation, and other mechanisms of post-transcriptional regulation. In addition, the 5’UTR overlaps the promoter region, and the corresponding DNA sequence can play a role in transcriptional regulation. Variation within the 3’ and 5’ UTR can result in functional consequences mediated through a variety of molecular mechanisms including, but not limited to, modification of RNA secondary structure, modulation of reading frame recognition by the translation machinery, and altered interaction with microRNAs (miRNAs) and RNA-binding proteins (RBPs).

As a direct result of the existence of clinically meaningful variation in UTRs, there is a need for accurate classification of pathogenicity for variants in these regions. Recent work has been conducted to develop recommendations for adapting existing variant classification guidelines to variants in non-coding contexts (<u>Ellingford et al., 2022</u>). Predictive algorithms have also been developed to assist in these efforts (<u>Moyon et al., 2022</u>; <u>Petrazzini et al., 2022</u>; <u>Wells et al., 2019</u>). However, the accuracy and consequent utility of such tools depends upon the quality of the data used for training and validation. Databases such as ClinVar (<u>Landrum et al., 2018</u>) and The Human Gene Mutation Database (HGMD) (<u>Stenson et al., 2003</u>) are frequently used as a source of training datasets. While valuable resources, conflicting classifications between submitters are common (<u>Rehm et al., 2015</u>) and frequent misclassifications have been documented (<u>Xiang et al., 2020</u>; <u>Shah et al., 2018</u>).

Given existing evidence of misclassification and known challenges of non-coding variant classification, we used ClinVar as a starting point for systematic curation and classification to develop a high-confidence set of pathogenic and likely pathogenic (P/LP) variants in the 3’ and 5’ UTR. Particular focus was placed on gathering mechanistic information, resulting in a collection of P/LP variants representing a range of proposed functional mechanisms. The census of P/LP variants presented in this work is further substantiated by findings from the application of deep learning (DL) models. These models demonstrated a distinction in prediction scores for variants acting through specific mechanisms relevant to the model, as compared to both P/LP variants acting through other mechanisms and putative benign variants. The resulting high-confidence set of curated variants will serve to support the continued development and accurate validation of DL algorithms designed to predict the functional impact of variants in the 3’ and 5’ UTR which may be implicated in rare disease.

## 2 Materials and methods

### 2.1 Variant identification and filtering

The tab delimited summary text file from the September 2022 release of ClinVar for GRCh38 was downloaded from the National Center for Biotechnology Information (NCBI) FTP site (last accessed October 20, 2022) (<u>Landrum et al., 2018</u>). Variants were further filtered to include only those with classifications of pathogenic, likely pathogenic, or with conflicting classifications of pathogenic and likely pathogenic across multiple submitters in ClinVar. Variants were annotated with allele frequencies from gnomAD v3.0 (<u>Chen et al., 2022</u>). Any variant with an allele frequency greater than 0.05 (5%) for any subpopulation was discarded, given this is sufficient evidence to warrant a benign classification (<u>Richards et al., 2015</u>). Variants were then filtered to include only those annotated in the transcript defined by ClinVar’s preferred name as 3’ or 5’ UTR exonic variants, defined as those located entirely within the coordinates of the respective UTR of the transcript, which must be coding. The transcript features were defined by NCBI RefSeq v109 annotations for *Homo sapiens* (<u>O’Leary et al., 2016</u>).

### 2.2 Curation of evidence related to variant effect and pathogenicity

The first step of the curation process consisted of confirming the variant’s 3/5’ UTR effect by considering the gene’s principal transcript as defined by APPRIS (<u>Rodriguez et al., 2022</u>). We excluded variants outside of the 3/5’ UTR of the most recent version of the APPRIS-defined principal transcript. In instances where a variant had a non-UTR effect on an overlapping protein-coding gene or non-coding RNA, the literature was consulted to determine whether pathogenicity was conferred through the UTR impact. If evidence suggested that pathogenicity was mediated by the non-UTR effect, or was insufficient to reach such a conclusion, the variant was excluded. Variants were also excluded in instances where they had a non-UTR impact on a *Matched Annotation from the NCBI and EMBL-EBI* (MANE) Select or Plus Clinical transcript for the same gene (<u>Morales et al., 2022</u>), regardless of having a UTR impact on the APPRIS-defined principal transcript. We also excluded variants with a non-UTR impact on an APPRIS-defined alternative or minor transcript when evidence implicating the variant as pathogenic was in the context of this transcript or there was insufficient evidence to discern the specific transcript mediating pathogenicity. Somatic variants, repeat expansions and structural variants with impacts extending beyond the UTR (i.e., large copy number variants (CNVs) impacting multiple gene regions or multiple genes) were excluded.

Synthesis of relevant evidence pertaining to each variant involved first consulting comments and reviewing citations provided directly in ClinVar. For each variant, an ancillary search was conducted to identify additional relevant literature. Data relevant to informing classifications of pathogenicity were extracted and documented. The information extracted depended on the nature of the study. For example, extracted details relevant to functional studies included the type of assay, results and overall conclusion. For case reports, the number of cases, their respective genotypes (e.g., homozygous, compound heterozygous) and phenotypic characteristics were recorded. Information related to proposed or validated mechanisms by which variants mediate their effects in the 3’ or 5’ UTR was captured, when available.

### 2.3 Classification of pathogenicity

Curated evidence informed classifications of pathogenicity based on guidelines published by the *American College of Medical Genetics and Genomics* (ACMG) and the *Association for Molecular Pathology* (AMP) (<u>Richards et al., 2015</u>). In keeping with recent recommendations for classifying non-coding variants (<u>Ellingford et al., 2022</u>), the following additional considerations were made: PS1 (strong criterion 1) was applied to variants impacting the same base as another variant previously established as pathogenic (e.g., a different change at the same base within a miRNA binding site). PM5 (moderate criterion 5) was applied for variants with the same predicted effect as a previously established pathogenic variant, but not at the same specific base/residue (e.g., a variant at a different site within a miRNA binding site in which another variant at a different base had been previously established as pathogenic).

### 2.4 Analysis of variant effects using deep learning models and conservation scores

To create the benchmark datasets for DL models, variants classified as P/LP were supplemented with putative benign variants obtained from gnomAD v3.0 (<u>Chen et al., 2022</u>). The datasets were created separately for the 5’ and 3’ UTR. For each transcript with a variant classified as P/LP in the corresponding UTR, the genomic coordinates of all exons within the respective UTR were used to construct an SQL query to extract variants within these regions from ‘bigquery-public-data.gnomAD.v3_genomes chr*’ tables on BigQuery. Variants were further filtered to have a total allele frequency greater than 0.01 (1%), a threshold more inclusive than that used for initial filtering of P/LP variants (0.05; see section 2.1), so as to allow for a greater likelihood of including a matched putative benign variant in the same UTR for the majority of P/LP variants, resulting in a more robust benchmark. Three models were applied to the resulting datasets: FramePoolCombined (<u>Karollus et al., 2021</u>), Saluki (<u>Agarwal and Kelley, 2022</u>), and Enformer (<u>Avsec et al., 2021</u>). Datasets were also annotated with PhyloP conservation scores (<u>Pollard et al., 2010</u>).

#### 2.4.1 FramePoolCombined predictions

The hg38 reference FASTA was downloaded from UCSC (http://hgdownload.cse.ucsc.edu/goldenPath/hg38/bigZips/hg38.fa.gz; last accessed Jan 11, 2023). A GTF file of the hg38 NCBI RefSeq table was downloaded from UCSC (http://hgdownload.soe.ucsc.edu/goldenPath/hg38/bigZips/genes/hg38.ncbiRefSeq.gtf.gz; last accessed Feb 15, 2023). A BED file of all 5’ UTR features was created from this GTF. Predictions for 5’ UTR variants were made using the Kipoi interface provided for the FramePoolCombined model (<u>Karollus et al., 2021</u>; <u>Avsec et al., 2019</u>). Each variant was predicted one at a time in its own VCF to yield individual variant effects, as the default behavior of the model through Kipoi is to integrate all variants in the VCF into the sequence for prediction. The predicted mean ribosome load (MRL) fold change was used as the variant effect prediction. Variants were stratified into three groups for analysis: “P/LP (open reading frame (ORF) mechanism)” if they were classified as P/LP and operated through a proposed mechanism of impacting ORF recognition by the translation machinery which included impact on an existing regulatory uORF or introduction of a novel upstream start codon (“model-matched”), “P/LP (Other)” for P/LP variants operating through a different or undetermined mechanism (“model-mismatched”), and “putative benign”. A Mann-Whitney Wilcoxon test was used to compare variant effect prediction scores between groups.

#### 2.4.2 Saluki predictions

For each 3’ UTR variant, the 6D track for the Saluki model, consisting of the one-hot encoded DNA sequence of the transcript, the coding frame, and the splice site positions was constructed for the reference and alternative sequence (<u>Agarwal and Kelley, 2022</u>). Predictions using all 50 cross-fold validation models provided by the authors were made for each of the reference and alternative sequences for the variant, and the average of the predictions from all models was used as the resulting score. The variant effect prediction was taken as the difference between the alternative and reference sequence scores. Variants were stratified into three groups for analysis: “P/LP (mRNA stability)” if they were classified as P/LP and proposed to impact the polyadenylation signal or mRNA stability (“model-matched”); “P/LP (Other)” for other P/LP variants operating through a different or undetermined mechanism (“model-mismatched”), and “putative benign”. The scores of the variants in each group were compared using the Mann-Whitney Wilcoxon test.

#### 2.4.3 Enformer predictions

Enformer was loaded via tfhub.dev (https://tfhub.dev/deepmind/enformer/1; last accessed Jan Feb 16, 2023). For each 5’ UTR variant, two sequences encompassing Enformer’s full context length were constructed: one centered at the reference allele and one centered at the alternative allele. For each sequence, predictions for the forward and reverse strand were made by Enformer, and the average was taken. The output was subset to the center window and two windows on either side (for a total of five windows) and only the CAGE tracks. The score for each sequence was calculated by summing over the five windows, adding a pseudocount of one, applying a log2 transformation, and finally computing the mean. The variant effect prediction was taken as the difference between the alternative and reference sequence scores. Variants were stratified into three groups: “P/LP (Transcription)” if they were classified as P/LP and operated at the level of transcription (“model-matched”); “P/LP (Other)” for all P/LP variants operating through a different or undetermined mechanism (“model-mismatched”), and “putative benign”. The scores of the variants in each group were compared using the Mann-Whitney Wilcoxon test.

#### 2.4.4 PhyloP conservation score annotations

The BigWig file of PhyloP conservation scores (hg38 100way, vertebrate alignments) was downloaded from UCSC (https://hgdownload.cse.ucsc.edu/goldenpath/hg38/phyloP100way/; last accessed Feb 22, 2023; <u>Pollard et al., 2010</u>). All 3’ and 5’ UTR variants were annotated based on the score at the corresponding position in the genome. In the event that a variant’s reference sequence spanned multiple nucleotides, the maximum score across the interval was taken.

## 3 Results

### Filtering and selection of 3’ and 5’ UTR variants from ClinVar

Variants documented in ClinVar were filtered based on classification (P/LP), gnomAD allele frequency (≤5%), and location within an exon of the 3’ or 5’ UTR (see section 2.1 for details). This filtering process yielded 295 and 188 ClinVar variants from the 3’ and 5’ UTR. Variants were assigned a confidence score as a means of initial assessment based on whether they were annotated to reside in regions outside of the 3’ or 5’ UTR on any other transcript (either an alternative transcript of the same gene or a transcript of a different gene). Higher scores reflect higher confidence that the variant’s effect is mediated through its impact on the 3’ or 5’ UTR of the ClinVar preferred transcript (Table 1). A score of two was assigned to 263 (89.2%) and 117 (62.2%) variants in the 3’ and 5’ UTR, respectively (Figure 1). All variants proceeded to curation irrespective of initial confidence score.

**Figure 1.**
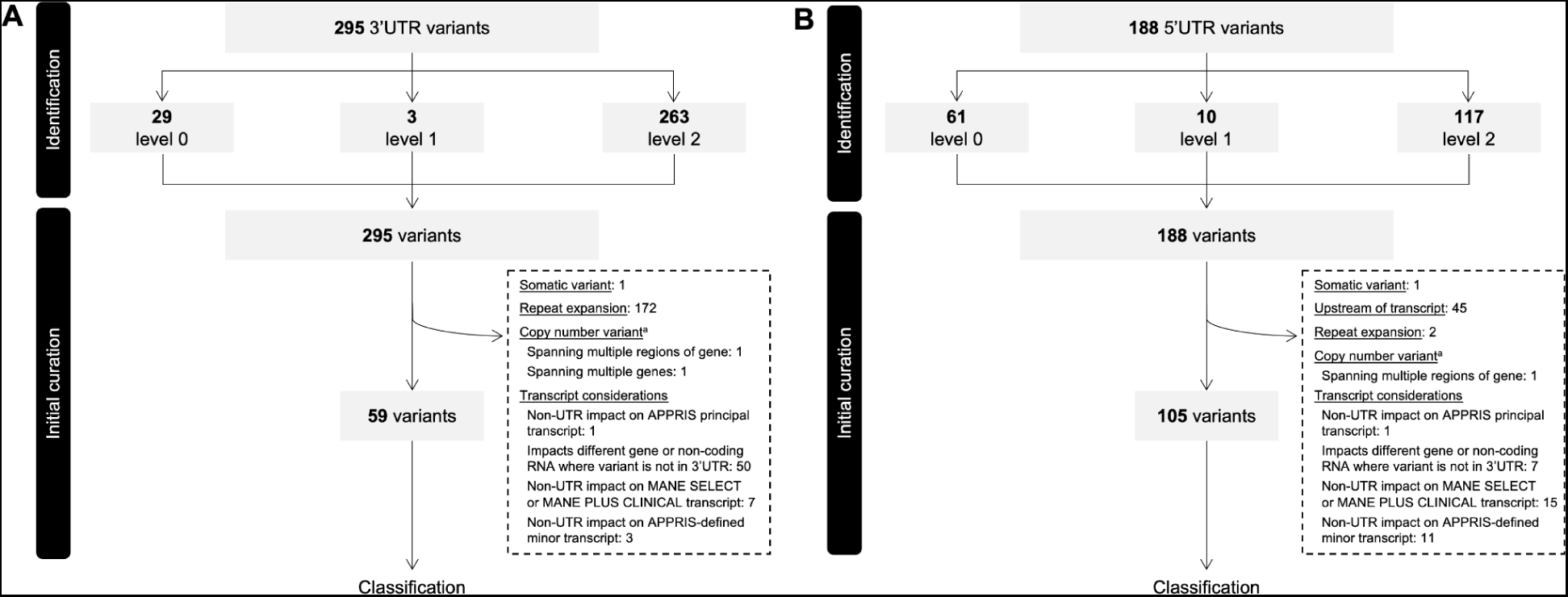
Flow of variants through identification and initial curation phases determining eligibility for classification. (A) 3’ UTR variants. (B) 5’ UTR variants. Variants were extracted from ClinVar, assigned an initial confidence level and filtered based on several factors before proceeding to classification. UTR, untranslated region; VUS, variant of uncertain significance ^a^Copy number variants include large insertions and deletions, exceeding 1,500 bp in length

**Table 1.** Initial confidence score assignment of variants based on ClinVar annotations.

|  |  |  |  |
| --- | --- | --- | --- |
| Criteria | Confidence Score |  |  |
|  | 0 | 1 | 2 |
| 3' (or 5' UTR) exonic variant based on ClinVar preferred transcript | ✓ | ✓ | ✓ |
| Not in a coding sequence exon in another transcript or a splicing variant<sup>a</sup> in another transcript | - | ✓ | ✓ |
| Not an intronic variant in another transcript or a 5' UTR (or 3' UTR) variant in another transcript | - | - | ✓ |

UTR, untranslated region
<sup>a</sup>Defined as an intronic variant within 8 bp of the intron/exon boundary

Of the initial 295 3’ UTR variants 59 (20.0%) proceeded to the classification phase, while the remainder (236 variants; 80.0%) were further filtered out for a variety of reasons, the most frequent of which being omission of repeat expansions (see section 2.2 for details; Figure 1A). All repeat expansions existed in *DMPK*, a gene associated with myotonic dystrophy 1 in which “normal alleles” consist of 5-34 CTG repeats within the 3’ UTR, typically asymptomatic “premutation alleles” between 35 and 49, and fully penetrant alleles over 50 CTG repeats (<u>Prior 2009</u>). Among the 172 *DMPK* repeat expansions in our dataset, 154 comprised >50 CTG repeats, while the remaining 31 involved between 31 and 50 repeats.

Of the initial 188 5’ UTR variants, 105 (55.9%) proceeded to classification. Among variants excluded, the most frequent reasons were transcript-related considerations (Figure 1B). One such consideration leading to variant exclusion was where, despite impacting a UTR of the APPRIS-defined principal transcript, variants had a non-UTR impact on an alternative transcript defined as either ‘Select’ or ‘Plus Clinical’ (i.e., transcripts not defined as ‘Select’, but in which known pathogenic variants have been reported) by MANE (<u>Morales et al., 2022</u>). Discordances of this nature were identified for seven 3’ UTR and 15 5’ UTR variants (Figure 1A-B). Three variants in *KRAS* serve as examples of the former, residing in the 3’ UTR of the APPRIS-defined principal transcript (NM_033360), but having missense impacts on the MANE Select transcript (NM_004985). *In vitro* functional evidence for one such variant (NM_004985:c.468C>G; NM_033360.4:c.*22C>G) as it resides on NM_004985 and induces a missense impact supports profound activation of the MAPK pathway (<u>Gremer et al., 2011</u>). In the absence of any such functional support for the variant as it resides in the 3’ UTR, its pathogenicity is more reasonably attributed to the missense impact. A collection of variants in *MOCS2* serve as examples of the latter, where despite residing in the 5’ UTR of the MANE Select and APPRIS principal transcript (NM_004531), non-UTR impacts exist on the transcript defined as MANE Plus Clinical (NM_176806). In these types of cases, and where there is no evidence providing mechanistic validation of any functional consequence of the variant in the context of a UTR of the APPRIS principal transcript, it remains challenging to ascertain whether any pathogenic impact is mediated by an effect on the UTR. These variants were therefore omitted from our dataset, in an effort to limit our collection of variants to strictly those for which we are confident pathogenicity is mediated through an impact specifically on the UTR.

Of the 59 3’ UTR and 105 5’ UTR variants proceeding to classification, the majority were classified as pathogenic in ClinVar (3’ UTR: 39 (66.1%); 5’ UTR: 72 (68.6%)), with the remainder classified as either likely pathogenic or a combination of pathogenic and likely pathogenic across multiple submitters (Figure 2A). Thirty-five and 54 unique genes were represented among the 3’ and 5’ UTR variants that proceeded to classification, respectively. The distribution of variants among genes is illustrated in <u>Figure S1</u>. Variants proceeding to the classification phase were classified according to guidelines published by the ACMG and AMP (<u>Richards et al., 2015</u>), with additional considerations made for variants in non-coding regions (<u>Ellingford et al., 2022</u>) (see section 2.3 for details). All variants, classifications of pathogenicity, and supporting evidence are documented in Tables S1 and S2 for the 3’ UTR and 5’ UTR, respectively.

**Figure 2.**
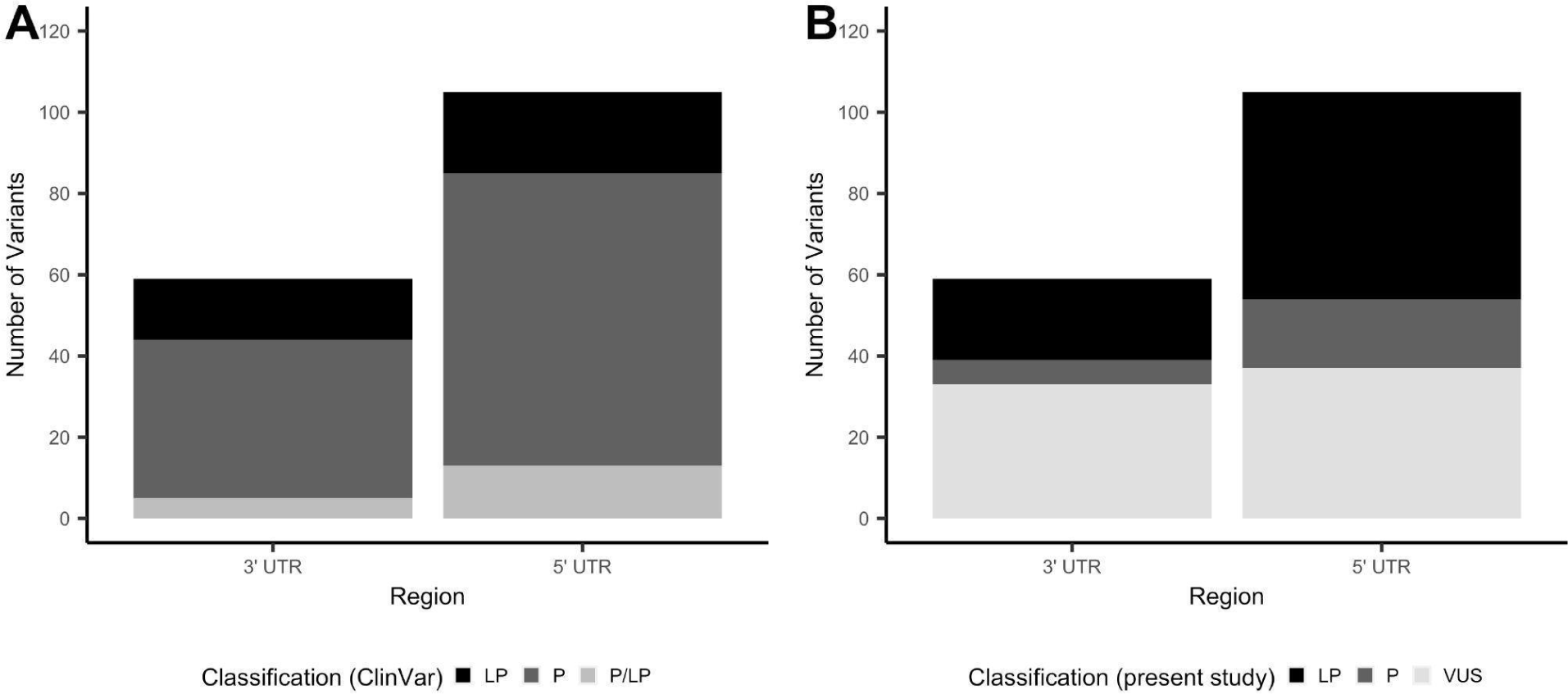
Number of 3’ and 5’ UTR variants by classification of clinical significance. (A) Total number of variants that proceeded to curation, stratified by clinical significance as reported by ClinVar submitters. (B) Total number of variants that proceeded to curation, stratified by clinical significance as classified in this study. LP, likely pathogenic; P, pathogenic; UTR, untranslated region; VUS, variant of uncertain significance

### 3.2 3’ UTR variant curation links pathogenic variants to a host of regulatory mechanisms

Of the 59 3’ UTR variants proceeding to classification, six were classified as pathogenic (10.2%), 20 likely pathogenic (33.9%) and 34 VUS (57.6%) (Figure 2B). 3’ UTR variants classified as pathogenic or likely pathogenic are provided in Table 2. Twenty-two of the 3’UTR variants classified as P/LP were supported by published functional evidence. All six variants classified as pathogenic in this work were also consistently classified as such by ClinVar submitters. Among the 20 variants classified as likely pathogenic in this work, 12 were pathogenic in ClinVar, four likely pathogenic, and four had conflicting classifications of pathogenic and likely pathogenic across multiple submitters.

**Table 2.**
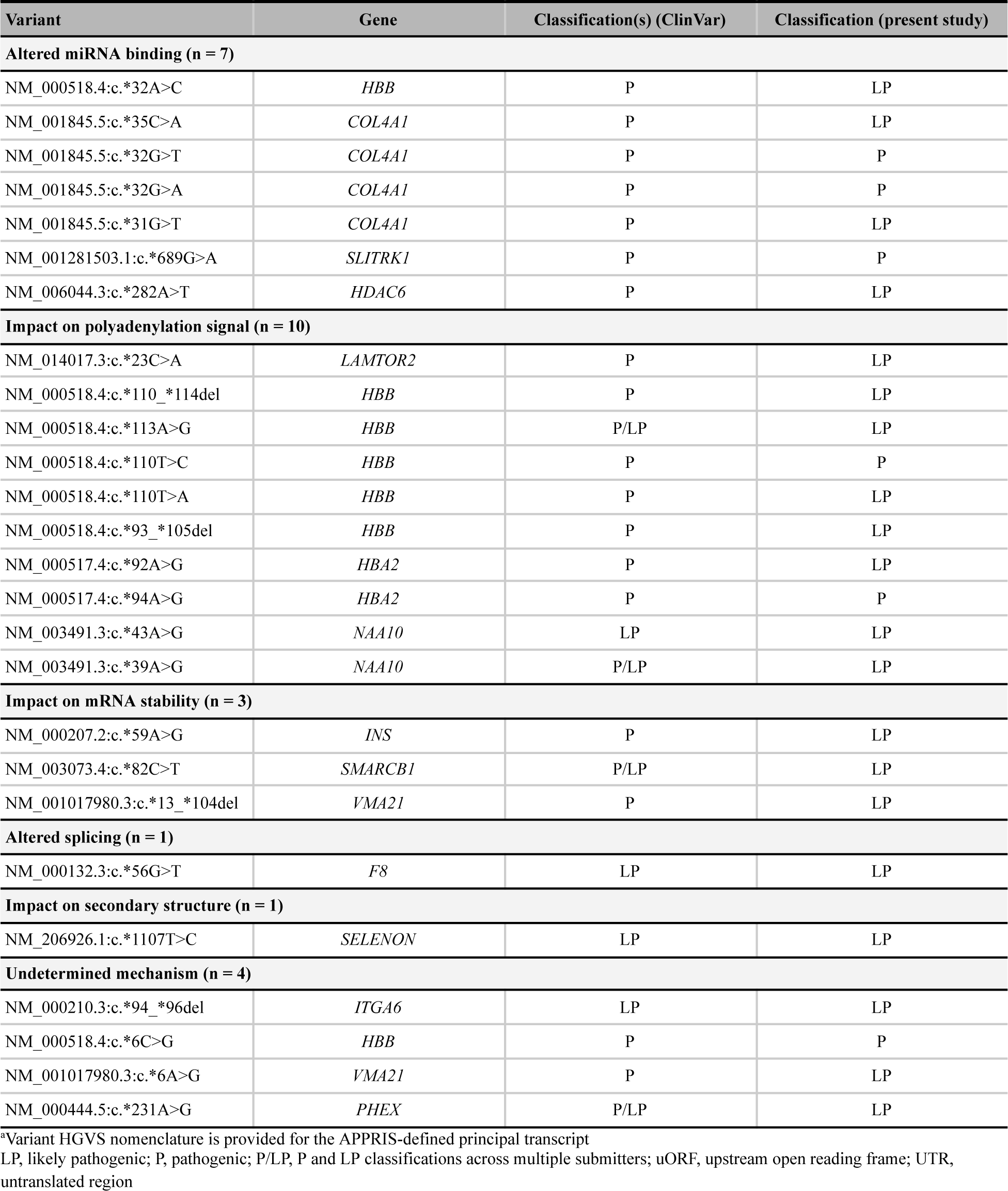
Pathogenic & likely pathogenic 3’ UTR variants by proposed or validated mechanism (*n* = 26)

|  |  |  |  |
| --- | --- | --- | --- |
| Variant | Gene | Classification(s) (ClinVar) | Classification (present study) |
| **Altered miRNA binding (n = 7)** |  |  |  |
| NM_000518.4:c.*32A>C | *HBB* | P | LP |
| NM_001845.5:c.*35C>A | *COL4A1* | P | LP |
| NM_001845.5:c.*32G>T | *COL4A1* | P | P |
| NM_001845.5:c.*32G>A | *COL4A1* | P | P |
| NM_001845.5:c.*31G>T | *COL4A1* | P | LP |
| NM_001281503.1:c.*689G>A | *SLITRK1* | P | P |
| NM_006044.3:c.*282A>T | *HDAC6* | P | LP |
| **Impact on polyadenylation signal (n = 10)** |  |  |  |
| NM_014017.3:c.*23C>A | *LAMTOR2* | P | LP |
| NM_000518.4:c.*110_*114del | *HBB* | P | LP |
| NM_000518.4:c.*113A>G | *HBB* | P/LP | LP |
| NM_000518.4:c.*110T>C | *HBB* | P | P |
| NM_000518.4:c.*110T>A | *HBB* | P | LP |
| NM_000518.4:c.*93_*105del | *HBB* | P | LP |
| NM_000517.4:c.*92A>G | *HBA2* | P | LP |
| NM_000517.4:c.*94A>G | *HBA2* | P | P |
| NM_003491.3:c.*43A>G | *NAA10* | LP | LP |
| NM_003491.3:c.*39A>G | *NAA10* | P/LP | LP |
| **Impact on mRNA stability (n = 3)** |  |  |  |
| NM_000207.2:c.*59A>G | *INS* | P | LP |
| NM_003073.4:c.*82C>T | *SMARCB1* | P/LP | LP |
| NM_001017980.3:c.*13_*104del | *VMA21* | P | LP |
| **Altered splicing (n = 1)** |  |  |  |
| NM_000132.3:c.*56G>T | *F8* | LP | LP |
| **Impact on secondary structure (n = 1)** |  |  |  |
| NM_206926.1:c.*1107T>C | *SELENON* | LP | LP |
| **Undetermined mechanism (n = 4)** |  |  |  |
| NM_000210.3:c.*94_*96del | *ITGA6* | LP | LP |
| NM_000518.4:c.*6C>G | *HBB* | P | P |
| NM_001017980.3:c.*6A>G | *VMA21* | P | LP |
| NM_000444.5:c.*231A>G | *PHEX* | P/LP | LP |

\*Variant HGVS nomenclature is provided for the APPRIS-defined principal transcript
LP, likely pathogenic; P, pathogenic; P/LP, P and LP classifications across multiple submitters; uORF, upstream open reading frame; UTR, untranslated region

A validated or proposed mechanism was reported for 22 variants, including disruption or introduction of a miRNA binding site (n = 7), impact on polyadenylation signal (n = 10), impact on mRNA stability (n = 3), introduction of a *de novo* splice site (n = 1), and change in secondary hairpin structure of the encoded mRNA (n = 1).

### 3.3 5’ UTR variant curation identifies variants spanning multiple pathogenic mechanisms

Of the 105 5’ UTR variants proceeding to classification, 17 (16.2%) were classified as pathogenic, 51 (48.6%) likely pathogenic, and 37 VUS (35.2%; Figure 2B). Fifty-five (80.9%) of the 68 variants classified as P/LP were supported by functional evidence. Of the 17 variants classified as pathogenic in this work, 12 were classified as pathogenic in ClinVar and three had a combination of pathogenic and likely pathogenic classifications across multiple submitters. Two pathogenic variants were classified as likely pathogenic in ClinVar, despite evidence cited in the respective submissions fulfilling sufficient criteria to warrant a pathogenic classification [<u>Richards et al., 2015</u>]. Across the 51 variants classified as likely pathogenic in this work, 38 were documented as pathogenic in ClinVar, six likely pathogenic, and seven pathogenic and likely pathogenic across multiple submitters (Table 3).

**Table 3.**
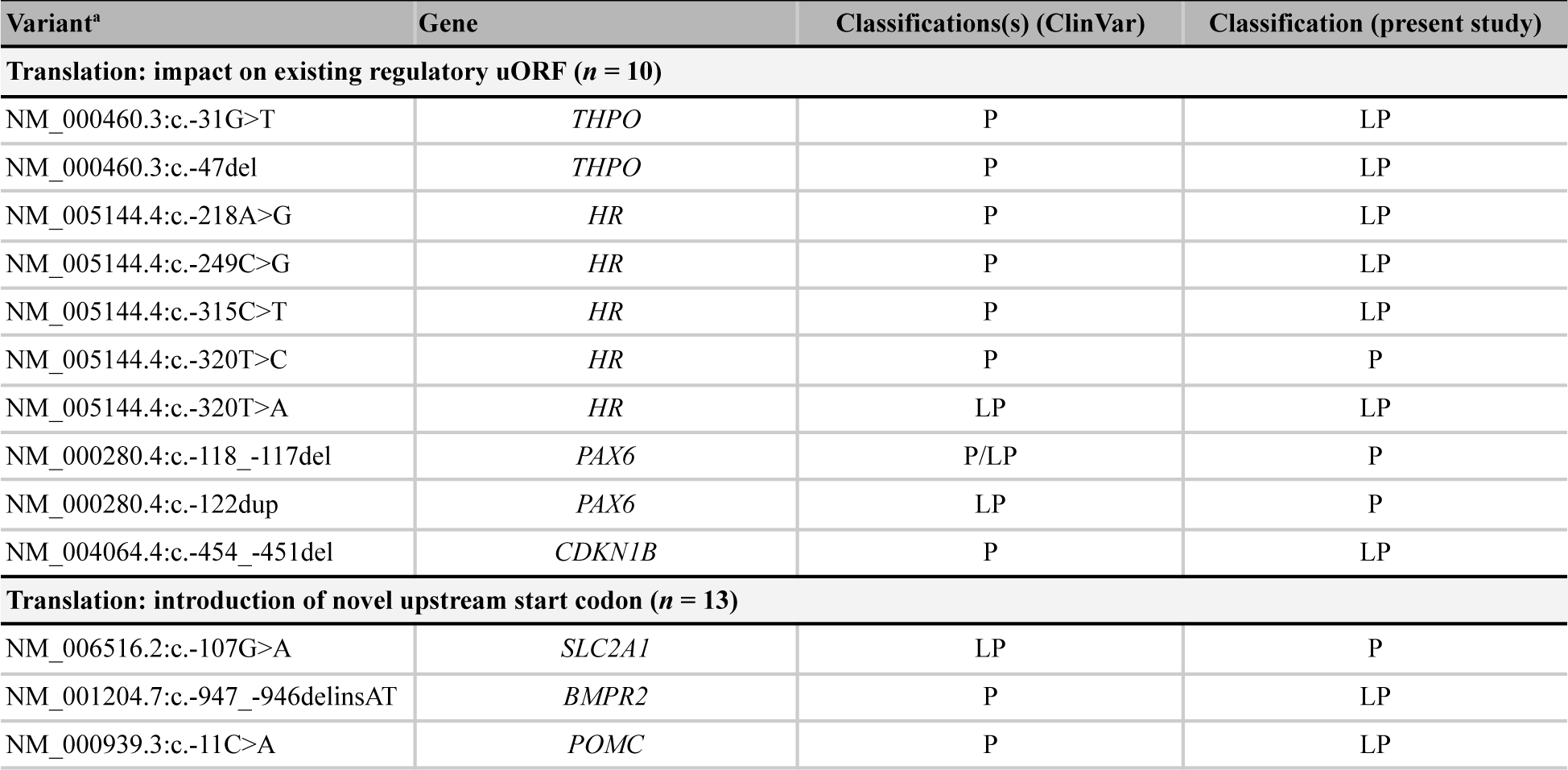
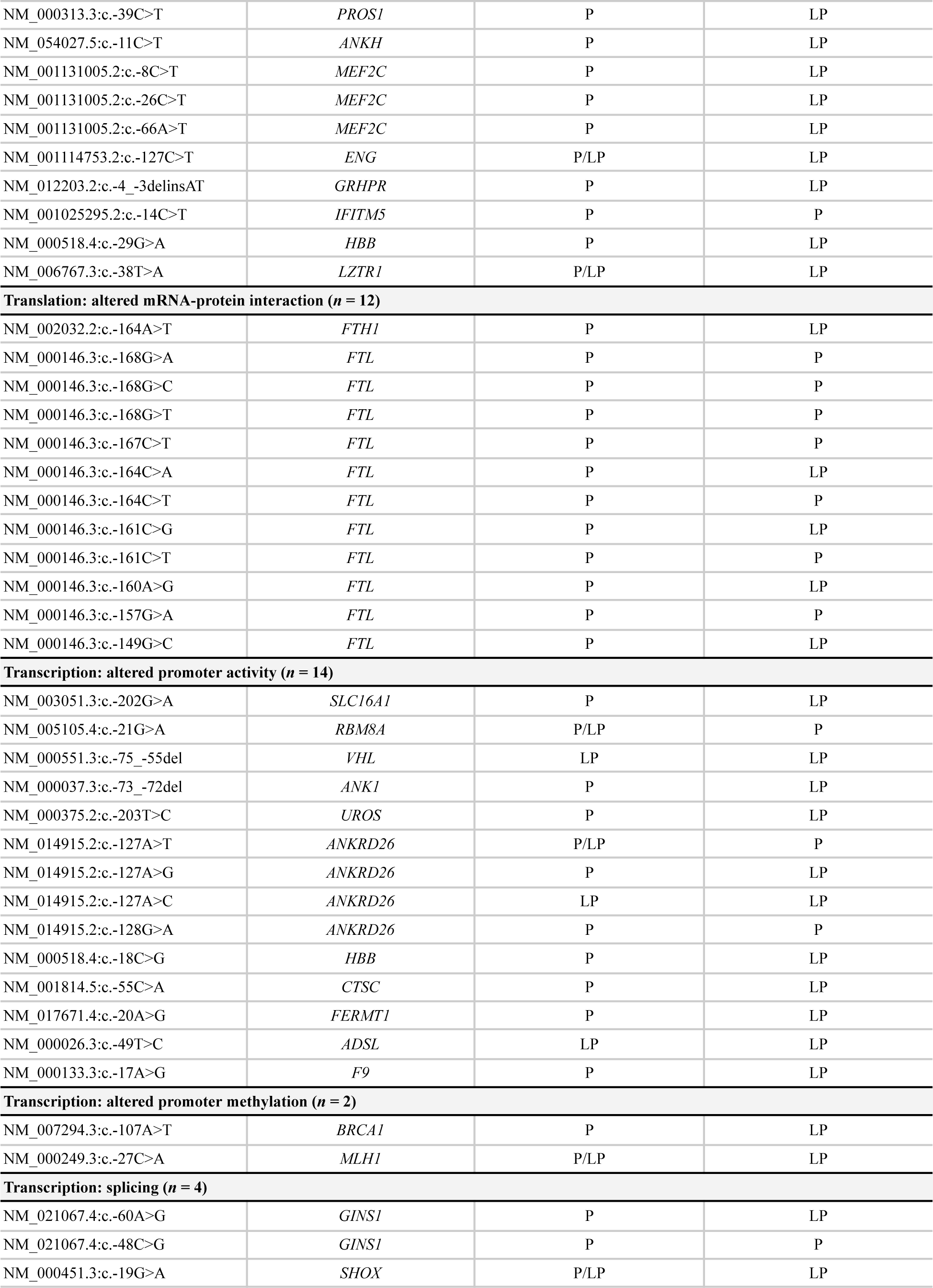
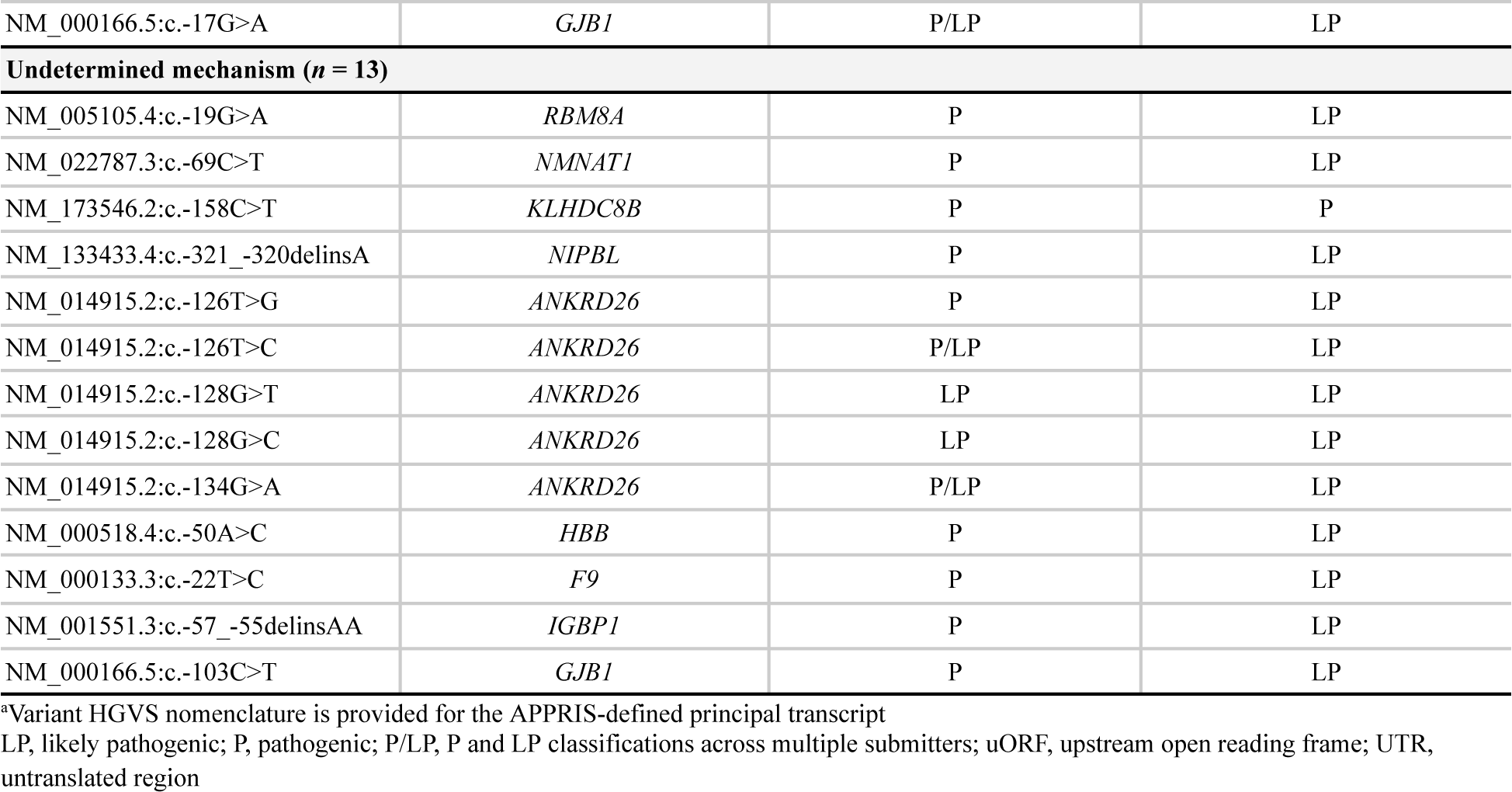
Pathogenic & likely pathogenic 5’ UTR variants by proposed or validated mechanism (*n* = 68)

A proposed or validated mechanism was also available for 55 variants classified as P/LP (Table 3). The described mechanisms operated both at the level of transcription (i.e., in cases where the 5’ UTR overlaps the functional promoter) and translation. Translational mechanisms included impacts on an existing regulatory upstream open reading frame (uORF; n = 10), introduction of a novel upstream start codon (n = 13), and altered interaction between mRNA and RNA-binding proteins (n = 12). Mechanisms operating at the level of transcription included altered promoter activity (e.g., enhancing or repressing transcription factor interactions; n = 14), altered promoter methylation (n = 2) and splicing impacts (n = 4).

### 3.4 Deep learning models capture the impact of curated pathogenic UTR variants

To further validate the curated set of P/LP 3’ and 5’ UTR variants, we investigated whether deep learning models trained for select UTR mechanisms could successfully capture the effects of the curated set of UTR variants in Tables 2 and 3. These models and their respective mechanisms are as follows: transcriptional effects in the 5’ UTR (Enformer; <u>Avsec et al., 2021</u>), impacting ORF recognition by the translation machinery (FramePoolCombined; <u>Karollus et al., 2021</u>), and mRNA stability in the 3’ UTR (Saluki; <u>Agarwal and Kelley, 2022</u>). For each of the 5’ and 3’ UTR, a dataset was constructed by selecting putative benign variants observed in gnomAD v3.0 that were in the corresponding UTR for any transcripts with a P/LP variant and had a total allele frequency above 1% (see section 2.4 for details; Tables S3 and S4). Though we tried to have at least one benign variant in the same UTR for every transcript, this was not always possible. This resulted in 68 P/LP variants and 24 putative benign variants in the 5’ UTR, and 26 P/LP and 67 putative benign variants in the 3’ UTR datasets.

Variant effect predictions were made by each model for the relevant UTR dataset. There was a significant difference (p-values ≤ 0.05 two-sided Mann-Whitney Wilcoxon test) between the absolute value of scores between all P/LP variants in their respective UTR combined and putative benign variants with all three DL models (<u>Figure S2</u>). A further delineation in scores was obtained when stratifying by mechanism, with model-matched P/LP variants having significantly higher scores when compared to model-mismatched P/LP variants (FramePoolCombined p-value ≤ 0.001; Saluki p-value ≤ 0.005; Enformer p-value ≤ 0.005), in addition to putative benign variants (Figure 3A-C). This finding highlights that the additional consideration of variant mechanisms is informative. Furthermore, considering the absolute value of model prediction scores was superior to considering the original predictions in differentiating model-matched, model-mismatched, and putative benign variants, particularly for Enformer (<u>Figure S3</u>). We anticipate that this may be due to variant pathogenicity being conferred through diverse mechanisms, including both expression increase and decrease. As such, the magnitude rather than the directionality of expression change, is more informative for stratifying pathogenic variants that operate through transcription as predicted by Enformer.

**Figure 3.**
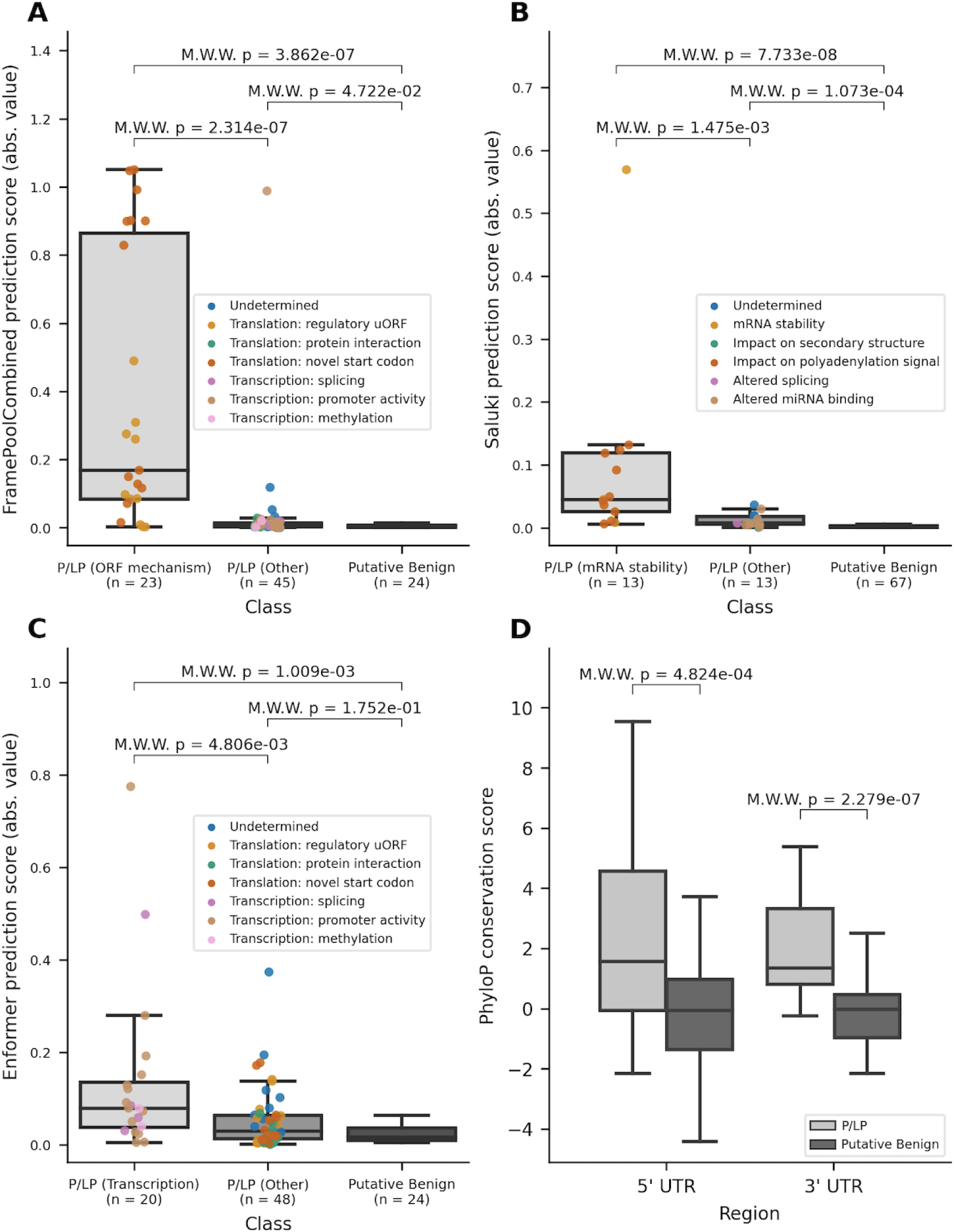
Variant effect scores from DL models, where the absolute value has been taken, and PhyloP conservation scores. (A) Distribution of the absolute values of scores for 5’ UTR variants as predicted by FramePoolCombined. (B) Distribution of the absolute values of scores for 3’ UTR variants as predicted by Saluki. (C) Distribution of the absolute values of scores for 5’ UTR variants as predicted by Enformer. (D) Distribution of PhyloP conservation scores for variants in the 5’ and 3’ UTR, stratified by pathogenicity. The boxplots have a center line for the median, the box limits are at the 25th and 75th percentiles, and the whiskers extend to 1.5x the 25th and 75th percentile values. Variants in the P/LP group have been colored based on their curated mechanism. M.W.W, Mann-Whitney Wilcoxon 2-sided test

The UTR datasets were then used as benchmarks to evaluate the performance of DL models on the binary classification task of differentiating between putative benign variants and model-matched P/LP variants. All models achieved greater than random performance, with area under the curve (AUC) values for the receiver operator curve (ROC) ranging from 0.66 to 0.84 (<u>Figure S4</u>). The performance was further improved when using the absolute values for all models’ scores, again suggesting that the magnitude of effect has greater predictive power for pathogenicity when applied to the select mechanisms for which deep learning models are trained to predict (<u>Figure S5</u>).

The availability of curated mechanistic data for this analysis provided valuable information for validating DL model predictions. For example, for P/LP variants that impact the ORF, variants introducing a novel upstream ORF (orange points; <u>Figure S3A</u>) tended to have lower MRL predictions from FramePoolCombined, likely due to the majority of novel uORFs resulting in out-of-frame translation (Table S2). In contrast, the two variants (NM_005144.5(HR):c.-218A>G, NM_000460.4(THPO):c.-47del) with the highest MRL predictions result in less inhibitory activity from an uORF, and greater translation of the main physiological ORF due to the disruption of an uORF, respectively. Further, there were nine P/LP variants in *ANKRD26*, of which eight were predicted by Enformer to result in increased expression (<u>Figure S3C</u>; Table S4). Though there was insufficient evidence to propose a definitive mechanism for all variants, curated evidence supported a mechanism in which variants likely result in loss of inhibitory regulatory action by RUNX1 and FLI1 (Table S2). This mechanism aligns with what would be expected of thrombocytopenia-associated *ANKRD26* variants, which have been documented to operate through a gain-of-function mechanism. (<u>Pippucci et al., 2011</u>).

Lastly, to supplement the support provided by the models, we also turned to conservation-based methods which have been used to identify potential regulatory regions in the non-coding genome before the recent advances of DL models. Each variant in the constructed datasets had its position annotated with PhyloP conservation scores (<u>Pollard et al., 2010</u>). There was a significant difference in scores between all P/LP variants and the putative benign variants within each of the UTR regions (Figure 3D), with PhyloP ≥ 1.6 offering the best separation between all P/LP and putative benign variants, when combining variants in both UTRs. To gain insight into the conservation of certain non-coding regulation mechanisms, we looked at the conservation scores stratified by mechanism. For variants in the 5’ UTR, variants that impacted translation tended to occur at nucleotide positions that were more conserved (<u>Figure S6A</u>). Variants that alter mRNA-protein interactions had the highest median PhyloP score, although this may be biased by the fact that all variants, except for one, are in FTL and occur in an iron response element (IRE), which is a conserved stem-loop (<u>Addess et al. 1997</u>). Variants that create novel upstream start codons were less conserved than those that impact existing uORFs, which is in line with existing findings that start and stop codons in existing uORFs tend to be highly conserved (<u>Whiffen et al., 2020</u>; <u>Lee et al., 2021</u>). Variants that affect transcription through altered promoter activity were located at sites that ranged between both high divergence and conservation, despite the importance of transcription factor binding motifs. For variants in the 3’ UTR, the ability to draw conclusions was limited by small sample size for most mechanisms. However, the mechanisms with the largest sample sizes, altered miRNA binding and polyadenylation signaling, both had median PhyloP scores greater than 1.5 (<u>Figure S6B</u>), consistent with these mechanisms relying on binding motifs and consensus sequences.

## 4 Discussion

In this study, our aim is to present a curated census of variants in the 3’ and 5’ UTR, with a focus on the mechanisms through which they confer pathogenicity. We curated a dataset of 26 and 68 variants classified as pathogenic or likely pathogenic in the 3’ and 5’ UTR, using a systematic approach and detailed evidence curation. Our analysis uncovered a considerable proportion of variants with interpretations of P/LP in ClinVar that lacked sufficient evidence to warrant such classifications. Reclassification of these variants highlights the need to exercise caution in utilizing public repositories without consideration of the level of evidence provided when evaluating variant pathogenicity. This finding is aligned with the ClinGen Sequence Variant Interpretation Working Group’s recommendation to omit “reputable source criteria” (i.e., PP5 (supporting criterion 5), BP6 (supporting benign criterion 6)) from clinical variant classification practices, citing the strong preference for primary data over expert opinion (<u>Biesecker et al., 2019</u>).

Beyond the variant sets themselves, our work provides insights into key challenges and considerations when classifying variants in non-coding regions. In their recommendations for adapting ACMG/AMP guidelines to non-coding variants, <u>Ellingford et al.</u> emphasize several important considerations, including classification in the context of the most clinically relevant transcript. Multiple methods have been developed to define biologically relevant transcripts, including the MANE collaboration’s approach, based on evidence including evolutionary conservation and expression level (<u>Morales et al., 2022</u>), and APPRIS, which defines principality based on cross-species conservation and information related to protein structure and function (<u>Rodriguez et al., 2022</u>). While we classified variants in the context of the APPRIS-defined principal, recent work by <u>Pozo and colleagues</u> reported a high level of concordance between MANE and APPRIS approaches, with agreement documented in over 94% of genes evaluated. In our experience, MANE Select and APPRIS principal transcripts were concordant in the majority of cases. We identified a few instances (3’ UTR: n = 7 variants; 5’ UTR: n = 15 variants) where, despite residing in the UTR of the APPRIS-defined principal transcript, variants had a non-UTR impact on a transcript defined as either ‘Select’ or ‘Plus Clinical’ (i.e., transcripts not defined as ‘Select’, but in which known pathogenic variants have been reported) by MANE (<u>Morales et al., 2022</u>). These findings highlight the importance of defining and classifying variants in the context of the most biologically relevant transcript, as classifications in alternative transcripts may yield different conclusions with respect to pathogenicity.

In addition to providing classifications as informed by applicable criteria, our datasets serve as a rich repository of mechanistic information accompanied by detailed summaries of functional evidence, where available. A diverse array of proposed mechanisms are represented across variants, operating at both the level of transcription and translation, albeit with varying levels of evidentiary support. Mechanistic information is relevant not only from the pathophysiological perspective, providing insight into functional underpinnings contributing to disease, but also with respect to the development and validation of mechanism-specific DL predictors of variant effects. Application of existing DL models to the curated 3’ and 5’ UTR datasets revealed a clear distinction in prediction scores when applied to variants operating through the mechanism for which the given predictor was designed. Furthermore, particularly in the case of Enformer, the absolute value of the scores was more informative in differentiating model-matched P/LP variants from the two other two groups. This finding reflects that pathogenicity can be conferred through both overexpression and lack of expression of a gene, as regulated through transcription. As such, the magnitude of change should be considered when evaluating the effect of multiple variants in aggregate, which may operate through both expression increase and decrease. Lastly, by combining PhyloP scores with this mechanistic information, we were able to provide insights into which non-coding mechanisms may be more greatly conserved in humans. Overall, our findings support the value of detailed and systematic curation of non-coding variant effects for the development, validation, and use of DL variant effect predictors in a mechanism-specific manner.

The datasets compiled and reported herein are not intended as an exhaustive resource of all P/LP 3’ and 5’ UTR variants documented in the literature, but rather a high-confidence set generated using a systematic approach consisting of both upstream bioinformatic processing and downstream curation and classification. Variants documented as P/LP in ClinVar were used as a reasonable starting point, although we recognize that non-coding variants are under-ascertained clinically and therefore under-reported in this database. Further, it is possible that true P/LP variants are currently documented in ClinVar as VUS, given historical challenges and lack of availability of classification guidelines tailored to variants in these regions. Increased implementation of WGS in the clinical setting and utilization of guidelines developed specifically for non-coding variants (<u>Ellingford et al., 2022</u>) will in theory increase the number of UTR variants documented as P/LP in ClinVar. These efforts can add to the breadth of the substrate available to contribute to the development of datasets similar to ours in the future. Also towards our effort to develop and implement a standardized and reproducible approach, we considered only variants for which a UTR impact was mediated through the variant’s effect on the APPRIS-defined principal transcript. It is therefore possible that variants with true pathogenic impacts mediated through UTRs on alternative transcripts, such as those residing within transcripts defined as MANE Plus Clinical, exist and are not represented in our datasets. Lastly, one limitation of our benchmarks for variant effect prediction is that not all P/LP variants could be matched to a putative benign variant in the same UTR of the same gene. As large-scale sequencing consortiums continue to focus on WGS, benchmarks for DL models can be improved upon to be more balanced and representative.

In conclusion, we present a high-confidence set of P/LP 3’ and 5’ UTR variants spanning a range of mechanisms and supported by detailed evidence curation and a systematic approach to classification. We anticipate these datasets to serve as valuable resources, given that variants documented in existing public repositories are not systematically curated in a consistent manner, and the reliability of such sources depends upon individual submitters. In generating this resource, we highlight key challenges and important considerations when conducting variant classification specifically in non-coding regions, including considering the potential for differential mechanistic impacts and classifications depending on the transcript considered. Findings from DL models further substantiate our classifications, with a distinction in scores supporting the relevance of mechanism-informed use of such predictors. These datasets will serve to support continued efforts towards developing and validating DL models designed to predict the impact of genetic variants residing in the 3’ and 5’ UTR.

## 6 Author Contributions

DM and TM conceived the study for this manuscript. EB, TL, OW, TM and DM contributed to developing the methodology. TL performed all upstream bioinformatic processing and conducted statistical analyses using DL models. EB curated supportive evidence and classified pathogenicity of all variants. TM supervised the study. EB and TL wrote the initial manuscript and created all figures. All authors reviewed and revised the manuscript.

## Data Availability

All relevant data are publicly available and provided in the supplementary material. Code to apply the deep learning models to the benchmarks and to produce all figures is available at the link.

https://github.com/deepgenomics/UTR_variants_DL_manuscript

## 8 Acknowledgements

We would like to thank Nasim Monfared for providing input on methodological aspects related to evidence curation, as well as Shreshth Gandhi and Albi Celaj for their input regarding the usage of deep learning models. We would also like to thank Brendan Frey, Laurence MacPhie, and Tom Peters for reviewing the manuscript for final approval.

## 10 Supplementary Material

**Figure S1.**
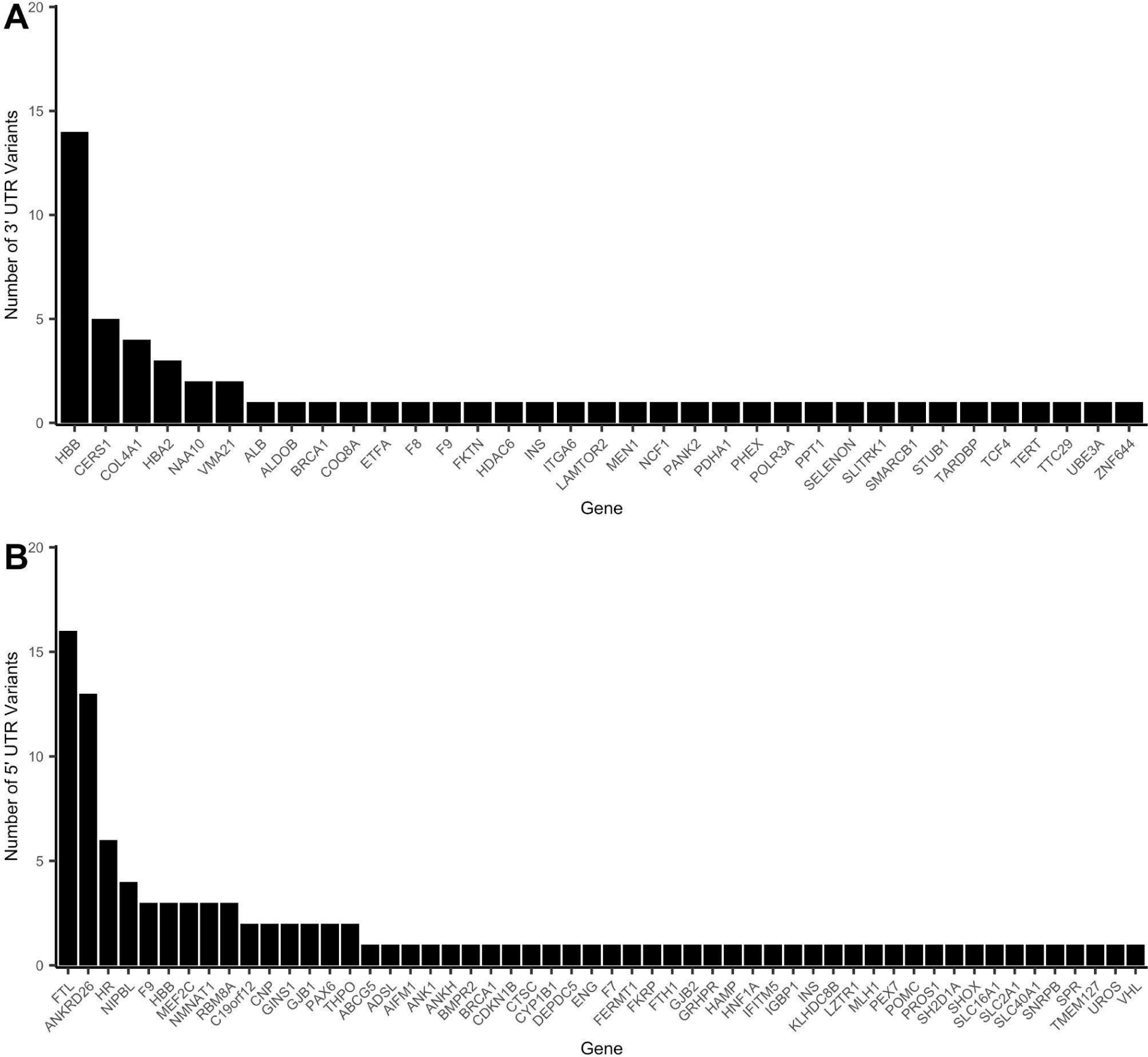
Distribution of variants across genes. (A) Genes represented across 61 3’ UTR variants proceeding to classification. (B) Genes represented across 109 5’ UTR variants proceeding to classification.

**Figure S2.**
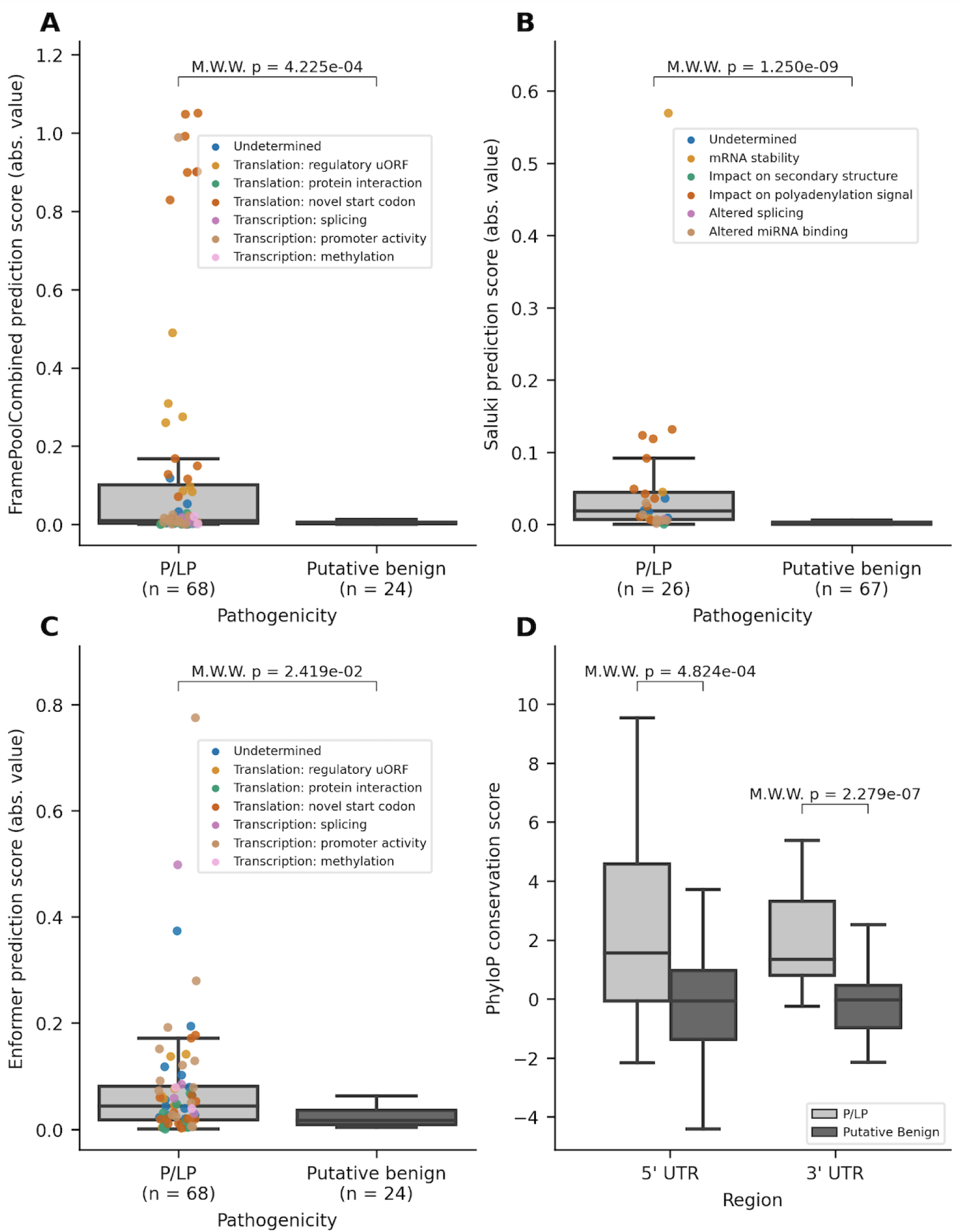
Variant effect scores from DL models (absolute values) and PhyloP conservation scores, with variants stratified by pathogenicity. (A) Distribution of the absolute value of scores for 5’ UTR variants as predicted by FramePoolCombined. (B) Distribution of the absolute value of scores for 3’ UTR variants as predicted by Saluki. (C) Distribution of the absolute value of scores for 5’ UTR variants as predicted by Enformer. (D) Distribution of PhyloP conservation scores for variants in the 5’ and 3’ UTR, stratified by pathogenicity. The boxplots have a center line for the median, the box limits are at the 25th and 75th percentiles, and the whiskers extend to 1.5x the 25th and 75th percentile values. Variants in the P/LP group have been colored based on their curated mechanism. M.W.W, Mann-Whitney Wilcoxon 2-sided test

**Figure S3.**
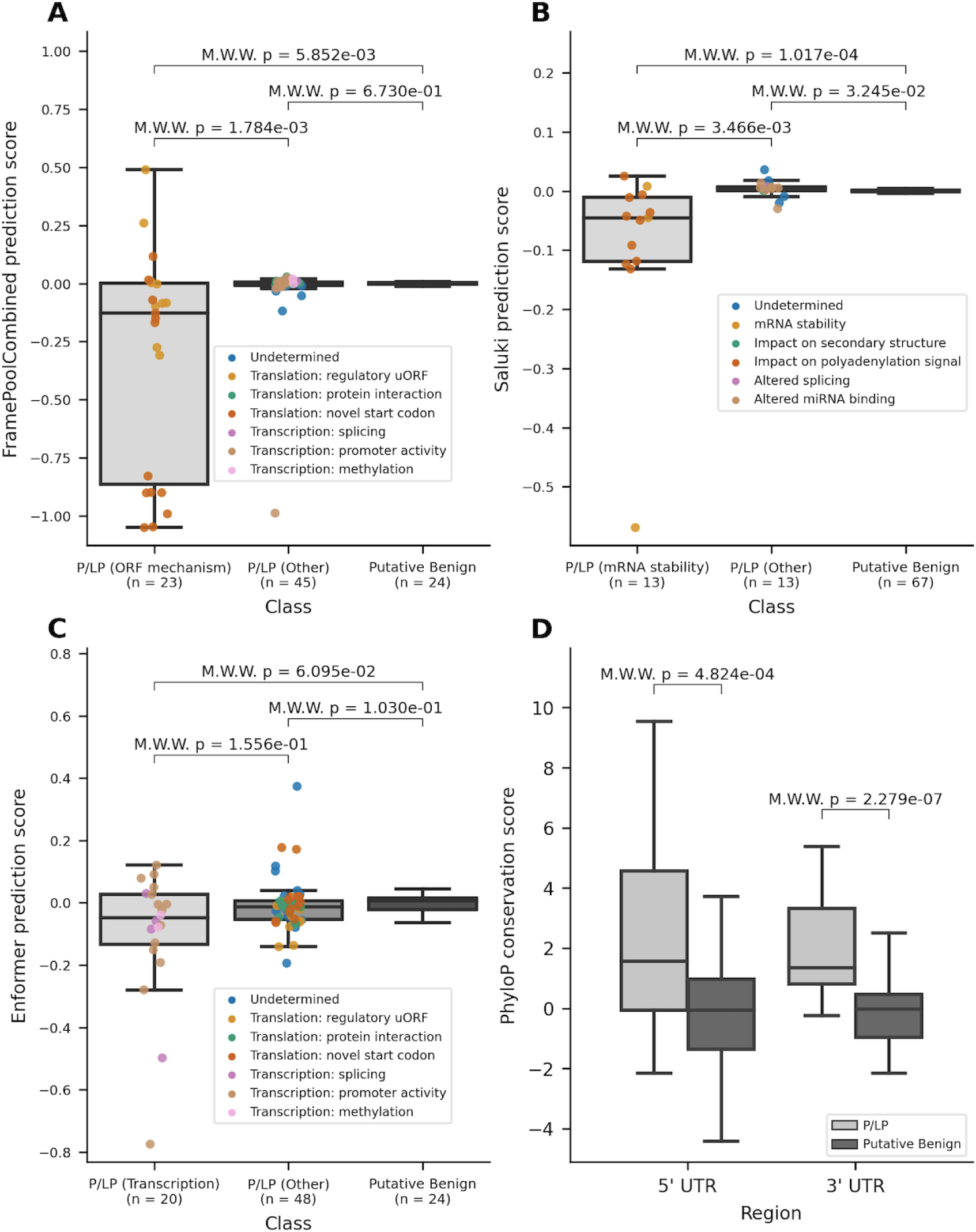
Variant effect scores from DL models and PhyloP conservation scores. (A) Distribution of scores for 5’ UTR variants as predicted by FramePoolCombined. (B) Distribution of scores for 3’ UTR variants as predicted by Saluki. (C) Distribution of scores for 5’ UTR variants as predicted by Enformer. (D) Distribution of PhyloP conservation scores for variants in the 5’ and 3’ UTR, stratified by pathogenicity. The boxplots have a center line for the median, the box limits are at the 25th and 75th percentiles, and the whiskers extend to 1.5x the 25th and 75th percentile values. Variants in the P/LP group have been colored based on their curated mechanism. M.W.W, Mann-Whitney Wilcoxon 2-sided test.

**Figure S4.**
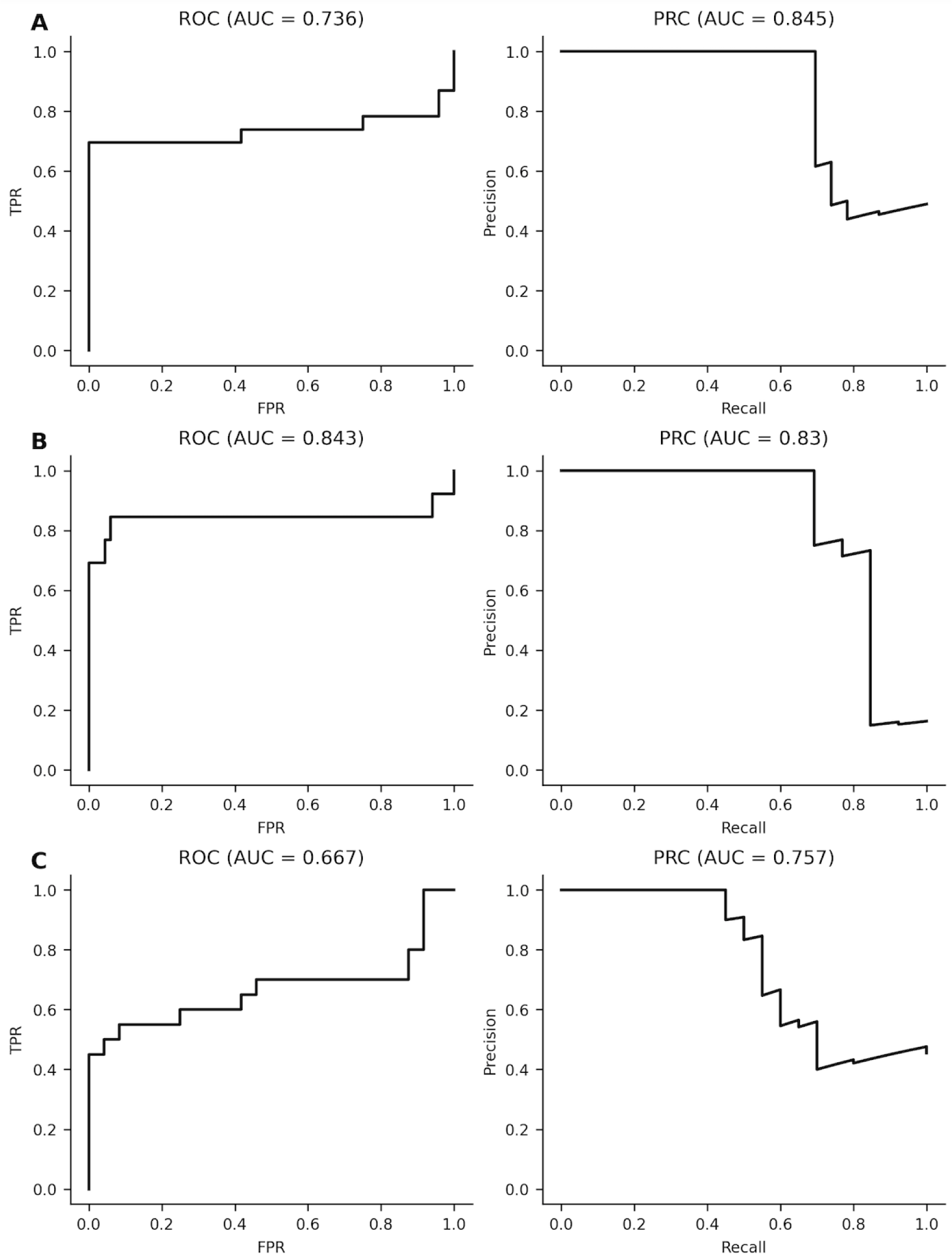
Receiver operator (ROC) and precision-recall (PRC) curves showing the performance of deep learning models in classifying model-matched P/LP variants and putative benign variants. (A) The performance of FramePoolCombined on classifying P/LP variants that affect the ORF (n = 23) and putative benign variants in the 5’ UTR (n = 24). (B) The performance of Saluki on classifying P/LP variants that affect mRNA stability (n = 13) and putative benign variants in the 3’ UTR (n = 67). (C) The performance of Enformer on classifying P/LP variants that affect transcription (n = 20) and putative benign variants in the 5’ UTR (n = 24) AUC, area under the curve; TPR, true positive rate; FPR, false positive rate

**Figure S5.**
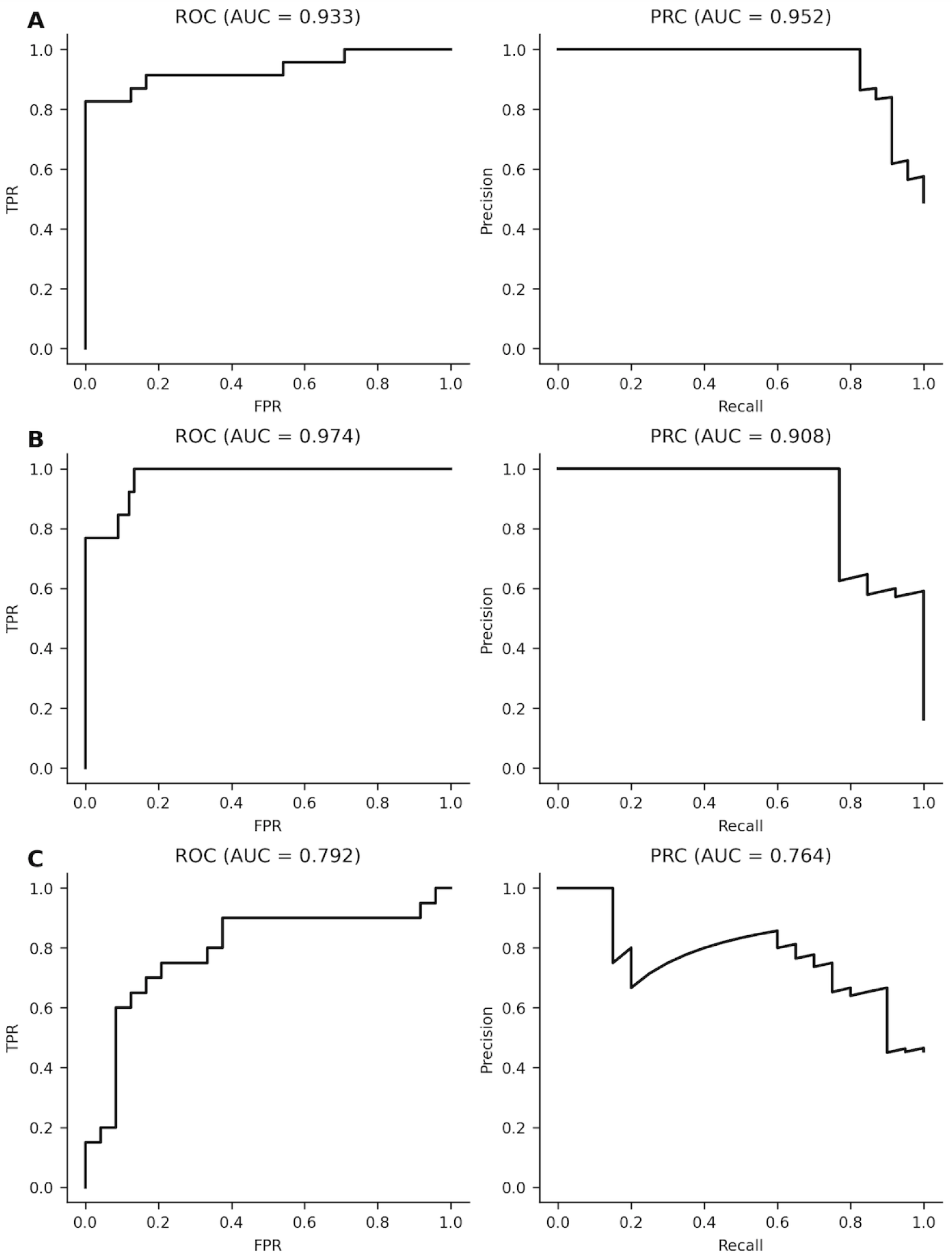
Receiver operator (ROC) and precision-recall (PRC) curves showing the performance of deep learning models in classifying model-matched P/LP variants and putative benign variants, using absolute values of the scores. (A) The performance of FramePoolCombined on classifying P/LP variants that affect the ORF (n = 23) and putative benign variants in the 5’ UTR (n = 24). (B) The performance of Saluki on classifying P/LP variants that affect mRNA stability (n = 13) and putative benign variants in the 3’ UTR (n = 67). (C) The performance of Enformer on classifying P/LP variants that affect transcription (n = 20) and putative benign variants in the 5’ UTR (n = 24) AUC, area under the curve; TPR, true positive rate, FPR, false positive rate.

**Figure S6.**
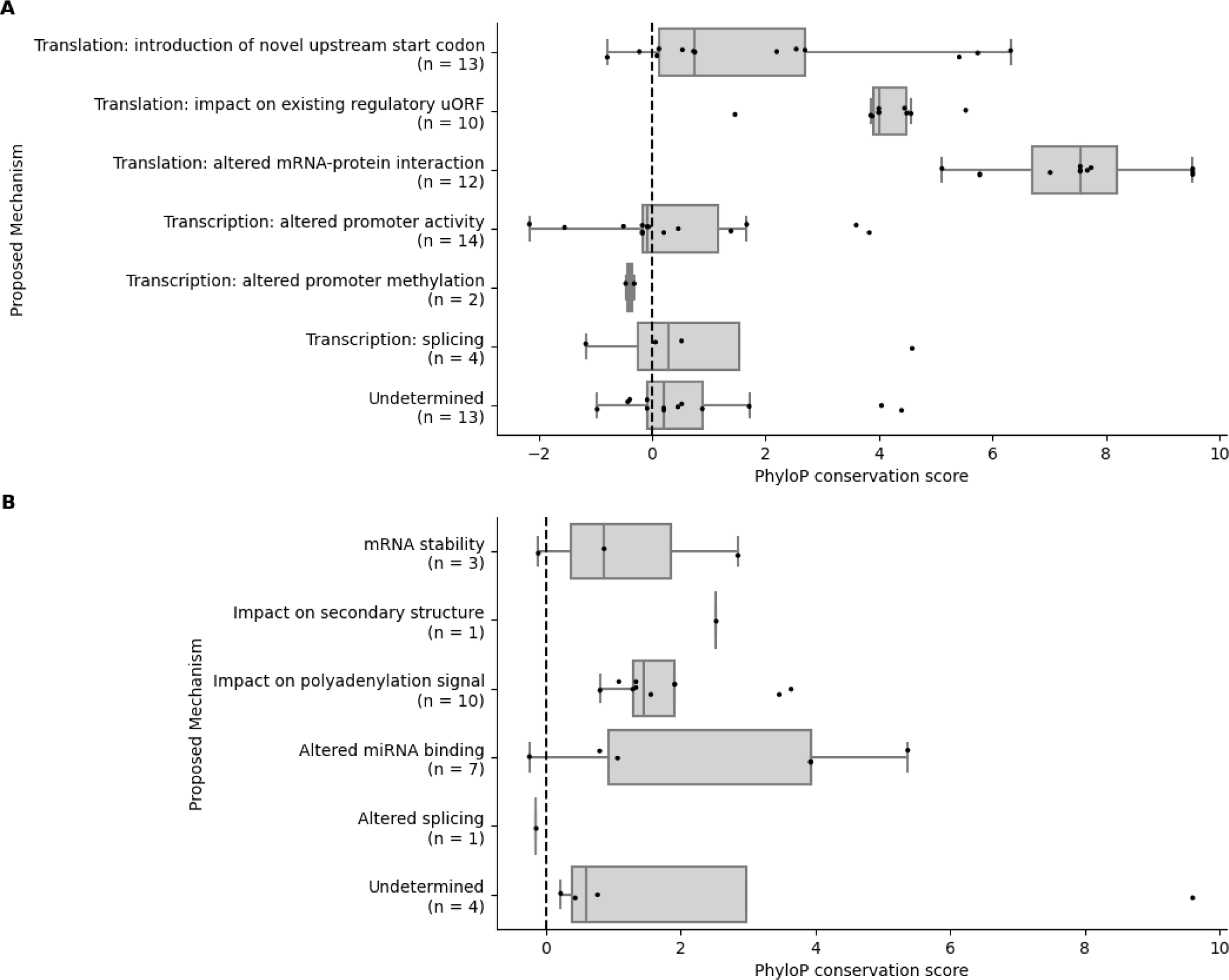
PhyloP conservation scores of P/LP variants, stratified by mechanism. (A) P/LP variants in the 5’UTR. (B) P/LP variants in the 3’ UTR. The boxplots have a center line for the median, the box limits are at the 25th and 75th percentiles, and the whiskers extend to 1.5x the 25th and 75th percentile values. The dashed line represents a PhyloP conservation score of 0.

**Table S1.** 3’ UTR variants: annotations, classifications & supporting evidence.

**Table S2.** 5’ UTR variants: annotations, classifications & supporting evidence.

**Table S3.** The 3’ UTR benchmark of pathogenic and likely pathogenic variants (hg38) and putative benign variants from gnomAD v3.0.

**Table S4.** The 5’ UTR benchmark of pathogenic and likely pathogenic variants (hg38) and putative benign variants from gnomAD v3.0.

## 11 Data Availability Statement

All relevant data are publicly available and provided in the supplementary material. Code to apply the deep learning models to the benchmarks and to produce all figures is available at: [https://github.com/deepgenomics/UTR_variants_DL_manuscript].

